# Pediatric intensive care hybrid-style clinical round during COVID-19 pandemic: A pilot study

**DOI:** 10.1101/2021.05.31.21258110

**Authors:** Mohamad-Hani Temsah, Ali Alhboob, Noura Abou Ammo, Ayman Al-Eyadhy, Fadi Aljamaan, Fahad Alsohime, Majed Alabdulhafid, Ahmad Ashry, Ahmad Bukhari, Omer ElTahir, Amr Jamal, Rabih Halwani, Khalid Alhasan, Jaffar A. Al-Tawfiq, Mazin Barry

## Abstract

**Background:** With the evolving COVID-19 pandemic and the emphasis on social distancing to decrease the spread of SARS-CoV-2 among healthcare workers (HCWs), our pediatric intensive care unit (PICU) piloted utilization of Zoom online into the clinical rounds to enhance communication among the treating team. We aimed to explore the feasibility of these hybrid virtual and physical clinical rounds for PICU patients from the HCWs’ perspective.

**Methods:** A mixed quantitative and qualitative deductive thematic content analysis of narrative responses from pediatric intensive care HCWs were analyzed, descriptive statistics were used

**Results:** A total of 31 HCW were included in the analysis; the mean time of the virtual round was 72.45 minutes vs. 34.68 for physical rounds, the most shared component in the virtual round was CXR (93.5%). Some of the HCWs’ perceived advantages of the hybrid rounds were enabling the multidisciplinary discussions, lesser round interruptions, and practicality of the virtual discussions. The perceived challenges were the difficulty of the bedside nurse to attend the virtual round, decreased teaching opportunities for the trainees, and decreased interactions among the team members, especially if the video streaming was not utilized.

**Conclusion:** Hybrid virtual and physical clinical rounds in PICU were perceived as feasible by HCWs. The virtual rounds decreased the physical contact between the HCWs, which could decrease the possibility of SARS-CoV-2 spread among the treating team. Still, several components of the hybrid round could be optimized to facilitate the virtual team-members’ interactions and enhance the teaching experience.

## Introduction

As SARS-CoV-2 infections continue to surge, with more than 140 million confirmed cases of COVID-19, including more than 2 million deaths, as reported to the WHO (1). With the second and third waves surging in multiple countries and several SARS-CoV-2 variants posing more challenges, the healthcare systems need more innovations to mitigate the surge in cases and protect the healthcare workers in the front lines (2, 3). Some health care systems emphasized the importance of infection prevention for the HCWs even outside the clinical areas due to their vital value (4).

Social distancing is one of the pillars of infection control measures, which may seem difficult to apply in the daily hospital rounds, where the whole healthcare team meets at the bedside, discussing the new clinical developments and best management approach for each patient. Previous research showed that most activities on attending rounds do not actually need to take place at the bedside(5). Another similarly important aspect of these bedside rounds is the clinical teaching and multidisciplinary interactions that are vital to the ongoing process of perfecting the healthcare professionals of the future with better utilization of healthcare resources which also does not necessarily require close gathering at the bedside (6).

After successfully implementing the virtual handover process of our pediatric intensive care unit (PICU) patients during the COVID-19 crisis (7), we planned to explore the feasibility of a hybrid morning daily clinical round in the pediatric intensive care unit (PICU) that was implemented since September 2020 (Appendix 1). This pilot study explores whether this hybrid round style decreased the timing of the physical proximity between the HCWs. Another aim was to facilitate the multidisciplinary team discussions, especially when several team members were not attending the hospital daily during the pandemic crisis.

## Method

### Study design

This study is a mixed quantitative and qualitative deductive thematic content analysis of the narrative responses from various HCWs in the PICU.

### Setting

The HCWs of the PICU at King Saud University Medical City (KSUMC) consists of six consultants, eight registrars, 4–6 training residents, two PICU fellows, 45 nurses, one pharmacist, one clinical dietician, one social worker, and rotating respiratory therapists. The PICU team serves 15 ventilated beds.

The hybrid rounds were newly implemented on May 15, 2020. Its structure consisted of starting the daily clinical round with Zoom^®^ meeting involving all the members of the PICU, including the physicians, bedside nurses, pharmacist, dietician, respiratory therapist, and social worker. The Zoom^®^ meeting was mainly devoted to discussing all the patients, including all the clinical data from all involved disciplines, suggested management plan, and discussing the educational aspect needed for specific clinical issues. After the virtual meeting, the on-call team, bedside nurses, and whoever is needed in the PICU will attend the physical rounds at the bedside for the issues requiring addressing there and counsel the parents about their child’s status.

### Sampling and recruitment

HCWs from various PICU backgrounds were invited to participate in this focused group on November 12, 2020.

### Data collection

Open-ended questions were used, as per Appendix 2.

### Data analysis

The first step in the analysis involved reading and familiarization with the participants’ range of responses. Categories were established, and two authors (NA and MT) developed codes independently. NA, an expert in qualitative methodology working in family and community medicine, introduced an etic perspective of the topic, while MT, a PICU consultant, introduced an emic perspective.

The developed codes were similar and were discussed before a consensus on the coding frame was established. All themes were a priori themes; however, the range of responses under each subtheme was derived from the data.

### Qualitative data management was conducted using NVivo 10^®^

After obtaining institutional review board approval, we invited PICU physicians of KSUMC, who have been performing hybrid using Zoom^®^, to describe their experience through a qualitative focused group virtual meeting.

Content analysis was used to analyze the participants’ responses. The results were used as a part of the quality improvement project and shared with the Pediatric Department Quality Committee.

### Statistical analysis

Descriptive statistics were used to summarize the data. For the categorical data, we used frequencies and descriptive procedures (minimum, maximum, mean, and S.D.). Bar charts give a graphical display of data using bars of different heights that are most frequently used for nominal and ordinal variables.

A line chart connects a series of data points with a continuous line was used to show the trend over time.

## Results

### Quantitative part

Our analysis has shown a clear difference between the time spent during the Zoom^®^ rounds compared to physical rounds, as shown in Table 1. Additionally, over the one-month pilot study period, the time spent during zoom rounds has dropped from 60 minutes in the begging to 40 minutes at the end of the month, although the number of patients was almost consistent throughout the month. This trend was also observed for the physical rounds, as time spent dropped from 38 to 18 minutes (Figure 1).

**Table 1:** PICU round Zoom time and physical round time (in minutes):

|  | N | Minimum | Maximum | Mean | Std. Deviation | Sum |
| --- | --- | --- | --- | --- | --- | --- |
| Zoom time | 31 | 45 | 122 | 72.45 | 22.590 | 2246 |
| Physical round time | 31 | 10 | 70 | 34.68 | 14.842 | 1075 |

**Figure 1:**
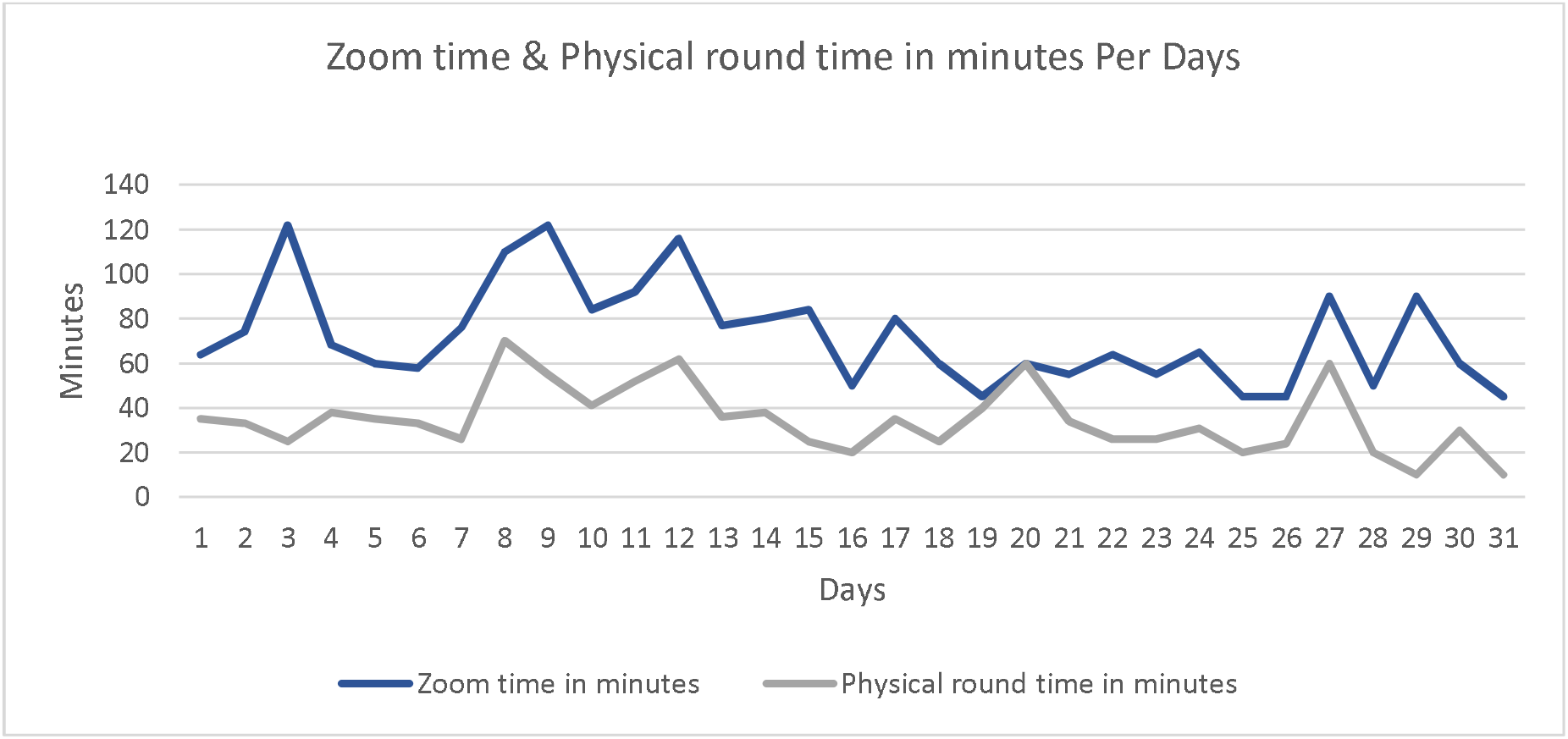
Zoom time & Physical round time in minutes per day.

Regarding the number of staff who attended the hybrid rounds, our results have shown that the number of HCWs attending the Zoom^®^ part has increased steadily over the study period, from 7 in the begging, to more than 15 at the end of the month, while the number of the staff who attended the physical (in-hospital) part remained somewhat stable over the study period (5-7), as shown in figure 2 and Table 2. The most commonly shared files during the Zoom meetings were chest X-rays, that were shared almost daily (Figure 3).

**Figure 2:**
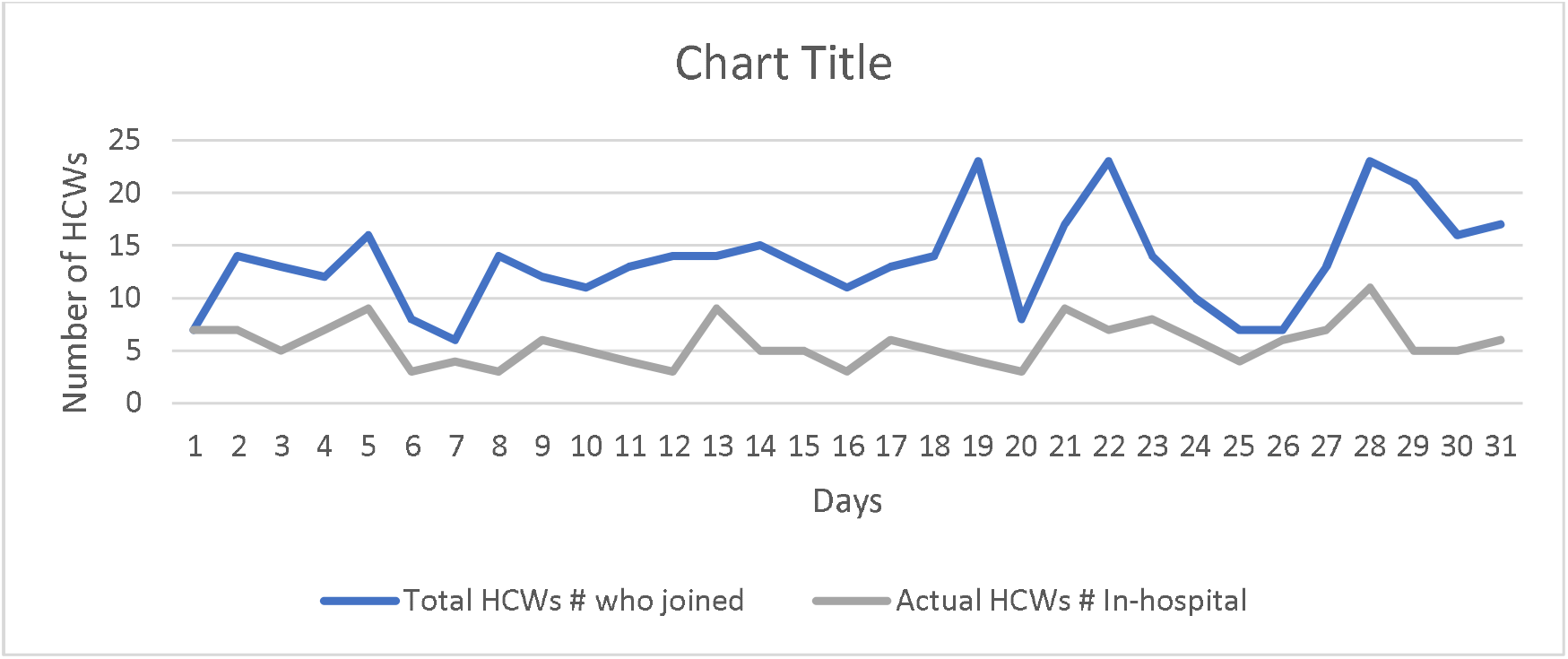
Total number HCWs who joined both the Zoom and the physical (In-hospital) part of hybrid rounds per day:

**Table 2:** HCWs who joined and actual HCWs from those in the hybrid round who are in-hospital:

|  | Minimum | Maximum | Mean | Std. Deviation |
| --- | --- | --- | --- | --- |
| Total number of HCWs who joined Zoom part of hybrid rounds | 6 | 23 | 14.29 | 4.224 |
| Actual In-hospital HCWs attending the physical part of hybrid rounds | 3 | 21 | 5.71 | 2.053 |

**Figure 3:**
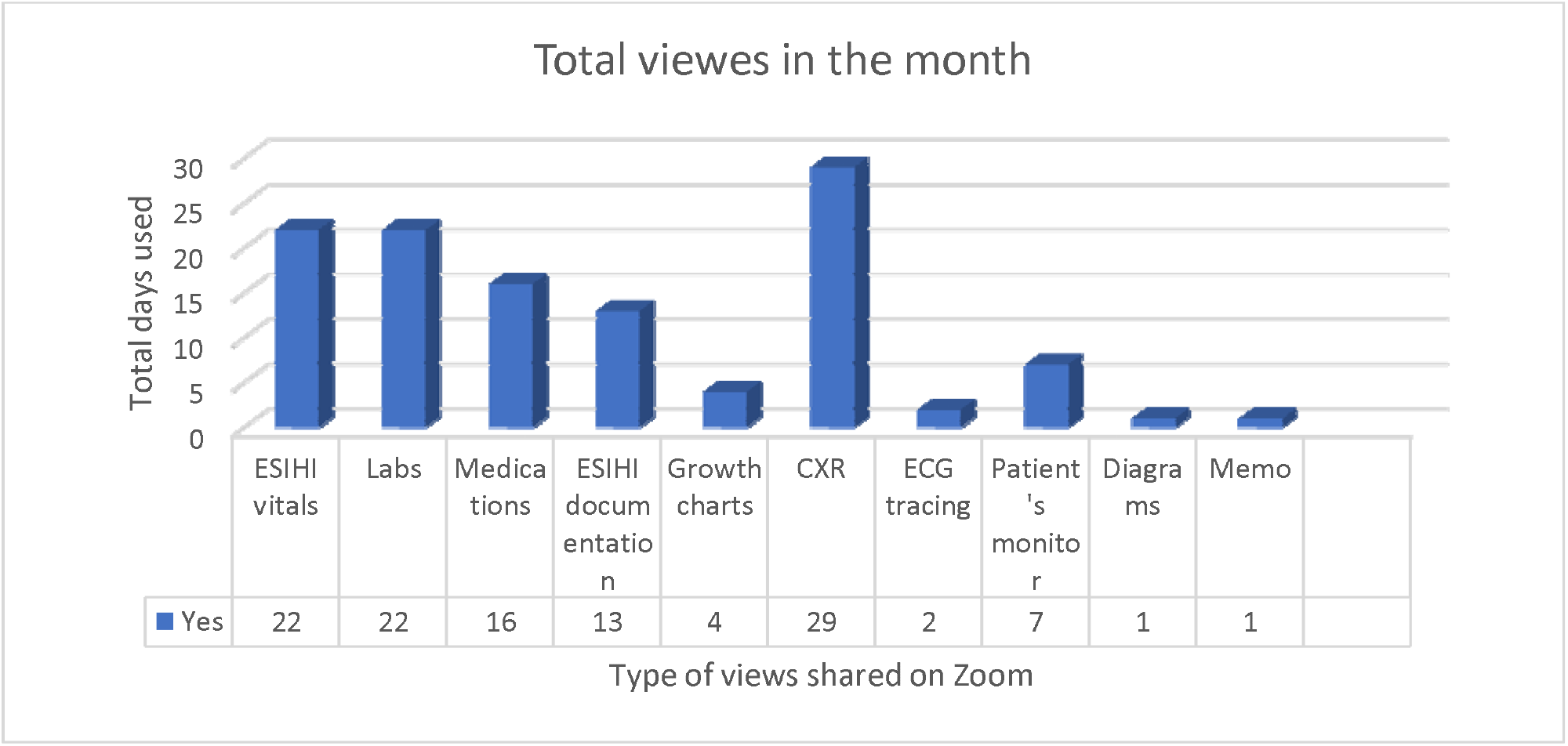
Total views per type shared on the Zoom round over the month. ^*^ ESIHI: Electronic System for Integrated Health Information, CXR: chest x-rays

### Qualitative part

Twelve PICU HCWs joined the focus group (F.G.): 3 consultants, three specialists, two training residents, two nurses, a pharmacist, and a dietician. During the meeting, participants discussed factors that affected their practice as a result of online rounds. The following presents the themes that were discussed during the F.G.

#### 1. Perceived advantages of hybrid round

Besides lowering the chances of being infected with COVID-19, participants mentioned other advantages they perceived using hybrid rounds. The following represents subthemes of perceived advantages.

##### 1.1. Multidisciplinary meeting

Participants believed that one of the most significant advantages of Zoom meetings is the opportunity to assemble people from different specialties at the same time to discuss patients’ conditions.

“The only thing that I think the zoom probably will be an advantage is for the multidisciplinary meetings regarding patients with different subspecialties joining. Otherwise, we do have difficulty arranging a meeting that suits everybody” P10

The participants appreciated the convenience of inviting colleagues from other disciplines to discuss PICU cases.

“you can invite any subspecialty, who could attend with us … if we need to discuss a specific patient for a specific concern” P1

##### 1.2. Family involvement

All participants appreciated the benefits they encountered regarding online communication between the PICU team and families. According to them, not having several family members at the bedside made them better focus on their clinical rounds, finish on time and give families undivided attention when discussing their children’s medical status.

One participant remarked:

“Now we focus more and avoid distractions from overcrowding areas … just avoid noisy areas with families gathering or interrupting the round” P8

Another one added:

“We have a special link for the families. Also, that is really very helpful for us because otherwise the families are coming during COVID crisis and interrupting the team dynamics, it is helpful for the PICU workflow.” P2

##### 1.3. The practicality of online meetings

Participants have mentioned many points related to the practicality of online rounds. All participants agreed that having online meetings from their offices was more convenient.

“it’s very convenient that you could be sitting all the time discussing things you could have your cup of tea or coffee in your office while you’re in the round” P10

They also mentioned being more efficient after the introduction of online rounds:

“My computer is in front of me, so I’m checking the patient during the Zoom round. I can check the labs, check the literature while they discuss the patient’s condition… When we finished the rounds, I can promptly put the orders in the EHR. Previously, with the physical rounds, I had to wait until we finish, and then I would go to my office and start doing the orders for the patients, which is more time consuming” P5

Another participant added:

“for us, Zoom meetings are effective: we can finish our task during the rounds. We can finish the orders swiftly” P11

#### 2. Perceived challenges of hybrid round

Participants in the focus group addressed some challenges that they faced during hybrid rounds. The following presents the subthemes that emerged from the discussion.

##### 2.1. Nursing duties

During the focus group, all nurses agreed on the difficulty of keeping up with online rounds and patient care at the same time. However, this is similar to the previous PICU rounds, as the nurses used to attend frequently to the patient’s needs, while the charge nurse would continue with the round. This was explained by one of the participants:

“the charge nurse should be present throughout the hybrid round… And the assigned nurse for each patient before the pandemic will be involved in the rounds when her assigned patient is being discussed. Now, if we are asking nurses to attend the whole Zoom rounds, that could be less time attending their patients?” P5

A head nurse added:

“Sometimes while we attend the Zoom round, something may happen to the patient, and the bedside nurse has to go inside the room immediately… so, when not discussing their cases, there is no point in attending the whole Zoom round” P3

The time nurses spent on online rounds was sometimes lengthy yet justified:

“It’s different from day-to-day, but it’s around one or one and a half hours. According to the number of patients we have and the severity of the cases and if they may need a lot of discussions or have multiple teaching points” P3

This challenge experienced by the nurses made some other participants look for a solution to overcome it. A resident commented:

“I suggest that we can have sequence numbering of patients that we are discussing. Number one would be the first patient to discuss, can be the sickest one. Number two, the less sick…and like that. The nurse would be able to know the order of when they are discussing her patient and be prepared to log in on time” P6

another consultant noted:

“So, if they can share with the nurses through one dedicated device, with the charge nurse handing it to each assignment nurse. So when we are discussing patient X, the device will be with the nurse assigned to patient X. They would attend for 15-20 minutes, then they will be focusing on patient care” P12

##### 2.2. Teaching opportunities

All residents within the focus group were concerned about learning and commented on the effect of hybrid rounds on teaching opportunities.

“Previously, there was more discussion and more teaching points to be addressed. I lost that sense in the hybrid rounds..” P11

One participant compared the discussions held at the bedside with those online, she explained:

“usually, bedside teachings were better. There’s more discussion and more brainstorming and motivates me…” P8

She continued:

“when things occurred during the round, the whole team is involved, to reflect on what happened … and how we manage it… This aspect of the teaching: we lost because we are sitting away from the patients during the round, just to be able to focus more and avoid all distractions” P8

Furthermore, residents commented on the effect of hybrid rounds on “X-ray rounds,” one participant explained:

“Previously, we used to start our PICU rounds with x-ray rounds, for 20 minutes discussing only all radiological exams of PICU patients. Nowadays, during COVID, we’re having Zoom rounds, we will show some x-rays, we will share the screen for the x-ray and sometimes pass it quickly.” P5

Participated consultants approved their residents’ concerns and explained their side of view on teaching using online methods. One of the consultants noted:

“Sometimes, I don’t feel motivated enough to teach during the rounds compared to the usual rounds. You can easily change your tone. You see the facial expressions you see who want to get more of your teaching. So that would all motivate you to give more” P10

Another consultant added:

“It (bedside rounds) used to be much more interactive with the application of knowledge rather than just didactic lecturing online or just answer the questions rather than building confidence into the solid background of the theoretical and practical approaches… people get “pulled away” from teaching if they are just facing a screen instead of reflecting on a patient look or patient monitor or ventilator or even something in the equipment has changed our approach to the medications. It’s (face-to-face teaching) totally much more engaging” P9

Furthermore, teaching was affected by the number of residents attending during the pandemic:

“During COVID, it’s really affected bedside teaching, especially for the residents. Daily, the hospital decreased the number of residents attending daily to enhance social distancing. Only three residents (from six or seven) attending inside the PICU. So, the clinical teaching is really compromised during this time” P12

Another participant added: “Still, some residents outside the hospital had the chance to participate in the Zoom round, so that could be a chance for more educational interactions even when they were not there in the physical rounds” P9

##### 2.3. Discussions and interactions

Although it was recommended that participants use their cameras during virtual rounds, they mostly did not:

“We tell people inside the hospital: please turn on your camera to make more interactions, especially for teaching or discussions. But mostly, they are not listening; they just open their mics when they decide to talk” P4

Proper engagement and interactions from the team members during the rounds were limited for reasons related to not seeing the speakers. A participant noted:

“the engagement and reading people body languages, getting people attention, focusing on what people need, reflecting on and building on ideas gradually as a group. This is usually done much better face-to-face, compared to just online …. The online style maybe limiting the team’s interactions” P9

A consultant noted:

“I think this sometimes may compromise a patient’s care if the nurse doesn’t speak up during the online meeting. During most of the online meetings, the nurses’ interaction seemed less compared, as to the face-to-face or physical rounds” P9

Another participant added:

“the Zoom becomes sometimes boring. Sorry to say that because, as with lectures, hearing is not like hearing and seeing, so you cannot fully concentrate and interact like when you are in physical rounds. You will also feel distracted since you are in your office; you can do other things while you are attending the round” P7

##### 2.4. The need to see patients in the PICU by all team members

Some participants thought that online meetings could not be blamed for compromising patients’ care, as explained in the following quotes:

“Physicians who are assigned with the patients, they are coming to the bedside” P3

“There are several physicians who are already available in the unit for the patients, so their care is not affected at all” P2

Other participants thought that they needed to see the patients when they need to. One participant illustrated:

“I go look at the patient and see if he/she is wasted or overweight and well-nourished. Now I miss those things because I don’t go to the PICU as before COVID. So, I depend on the numbers, like height and weight, which are sometimes not accurate, so I must check with the doctors and nurses about the patient’s physical appearance.” P5

##### 2.5. Role of R.T.s in the online rounds

Despite participants’ acknowledgment of the convenience of multidisciplinary meetings using online methods, this did not facilitate involving R.T.s, whose roles were perceived as crucial for patient care quality in the PICU. One of the consultants clarified:

“before we are having dedicated R.T. to the PICU. Yeah. Now, during the COVID crisis, the R.T.s are shared with two or three other units, there is more demand for their services during the pandemic. It’s mainly a respiratory pandemic. So that’s a problem… it was another challenge to invite them because the R.T. has very valid points to raise when we talk about mechanical ventilation. We need them for the full respiratory management of these children” P9

## Discussion

Close contact with SARS-COV-2 carriers becomes riskier as more time is spent in such encounters. The Center for Disease Control and Prevention (CDC) reported in its Morbidity and Mortality Weekly Report (MMWR) on April 14, 2020, about HCWs who developed COVID-19 with having longer durations of exposure to the index COVID-19 patient (8). These findings underscore the heightened COVID-19 transmission risk associated with prolonged, unprotected patient contact. The CDC recently reported that the recurring brief encounters could lead to COVID-19 transmission (8). This could be an additional risk for the HCWs in the acute care areas who manage COVID-19 suspected patients. Furthermore, asymptomatic SARS-COV-2 infection HCWs was reported in 14.3%, implying that strict infection measures among HCWs to reduce transmission risks to other HCWs and patients (9).

The implementation of the Zoom hybrid round was feasible yet challenging in our PICU setting. The utilization of technology as novel communication tools during the pandemic and Tele-ICU was advocated in recent literature (7, 10, 11). During times of physical distancing, HCWs may find it helpful to use videoconferencing services to sustain professional meetings and continue educational activities using online platforms (10). Recent policy changes in telemedicine during the COVID-19 pandemic have generated technology-based clinical tools opportunities, which could help conserve personal protective equipment (PPE) and protect HCWs (12). Such a new approach was labeled as electronic PPE (ePPE), which would maintain clinical services, preserve the actual PPE, and keep HCWs safe (12).

Virtual rounds in our pilot period replaced 60% of the physical round. The time saved was utilized to enhance and augment the discussion about the patients’ condition, laboratory findings, and teaching. Such rounds are essential aspects of the education and teaching of medical students, interns, and residents, allowing them to understand key information and develop clinical reasoning(13). Virtual rounds may also decrease the embarrassment that students may have due to the presence of patients. However, one drawback is the lack of patients-learners interaction(13, 14). Other studies showed that virtual rounds specifically designed to manage COVID-19 patients had a favorable encounter by patients and learners (15). It had also been shown that virtual teaching is effective and may further enhance education by the availability of different specialties (16).

### Our hybrid clinical round setting

allowed multidisciplinary team’s, reaching up to 23 HCW at the same time while maintaining social distancing. In a perspective piece on remote pediatrics health care delivery during the pandemic, researchers highlighted the different variations and innovations in adaptation and called for integrating telemedicine and virtual health (17). Our hybrid approach can be adapted and validated by other PICUs, where the number of HCWs may exceed ours, and when physical distancing may not be feasible. Although vaccination of HCWs is being rolled out with excellent efficacy data (18, 19), the emergence of mutant variants(20-22) would continue to challenge HCWs and would still imply continued universal masking and physical distancing(23).

Our study participants commented about the need for flexibility in the hybrid round to allow for patient care as needed. ICU nursing staff have been under unprecedented pressure during the pandemic and showed resilience and continued patient care despite all stresses and pressure (24); this type of hybrid rounds would help alleviate some of that pressure without compromising patient care.

Some overwhelmed clinical services, like R.T.s, were unable to join the Zoom part of the rounds in our setting. With the overwhelming number of critical care patients requiring respiratory support, a surge in demand for respiratory therapists was seen, with safety, treatment, and staffing recommendations published(25), and evaluating tools on R.T. extended comfort with mechanical ventilation during the pandemic, a hybrid type of round may help alleviate such pressure on R.T.s allowing them to have more time to tend to their patients (26).

This study showed that the most shared items during the virtual clinical round were radiography, EHR vitals, and laboratory data. The ability to share radiographs is an essential aspect of the Zoom clinical rounds. In one study, radiology residents could transition the teaching conference and educational lectures entirely to a virtual format (27). Of course, as indicated previously, one possible extension of this feature is that trainees may seek input from more senior physicians using the Zoom methods (27). This feature was highly appreciated by the residents who were involved in our study. In addition, it is very convenient to read out, or share screens to discuss other patients’ related data such as laboratory data and vital signs either as data points or utilizing any graphic presentations, provided the patient’s confidentiality is maintained with Zoom’s end-to-end encryption (28).

Although telehealth has multiple advantages, it also has its pitfalls. One such pitfall is that physical examination might not be optimal, especially for the new physicians. In one study, telemedicine demonstrated poor agreement with an in-person examination of patients with tonsilitis (29). Few studies are addressing the validity of virtual examinations (30). Since it is likely that telemedicine for examination will continue beyond the pandemic, some studies are looking at proposals for such examinations (31). This requires additional skills of the individuals using telemedicine, including robust communication skills and the ability to perform such examinations remotely and accurately.

Participants suggesting that the clinical teaching for the training residents was really compromised during COVID-19, especially for those outside the hospital. The Zoom round seemed to have both negative and positive impacts on the teachings but needs to be optimized. It is crucial to build telemedicine into the residency program’s curriculum and medical students to expose them to the advantages and disadvantages of telemedicine (32). In addition, it is essential to address technical proficiency, history taking, examination skills, and communication (33). In addition, it is important to combine both telemedicine and in-person clinical rounds as a hybrid activity. A pilot study of the use of telemedicine in primary care showed general acceptability with logistics and other concerns. It is important to note that the exposure to telemedicine by medical students and 17.4% of students had prior telemedicine exposure (34). Such integration of telemedicine into medical education is of paramount importance (35).

The presence of the family during rounds and sharing decisions will increase family satisfaction during admission and may enhance the communication between the treating team and parents(36). However, frequent parental interruptions of the PICU round will increase the round time and affect the team dynamics(36, 37). So hybrid round started initially before in-person which can gather information about the patient and sharing decisions among medical team providing teaching opportunity for the rotators without interruption from the family(38). Still, the possibility of a decrease in the quality of face-to-face interactions between the residents could affect the interactive family scenarios in the PICU (39).

## Limitations

This single-center experience needs to be validated in other settings. Training may be more challenging in other hospitals, as our PICU team was already using the videoconferencing by Zoom for patients’ handover. While we did notice a decrease in the time of the hybrid round over the one-month study period for the same number of PICU patients, this observation might still point to the team’s learning curve, which has improved over time with the new hybrid system of daily rounds. However, this study was not designed to examine the HCWs’ learning curve, and reporting learning curves in health professions research is deficient and often underutilizes their desired properties (40).

## Conclusion

Hybrid-style rounds in the PICU were feasible and decreased the timing of the physical round. The virtual component of the daily hospital rounds facilitated multidisciplinary discussions and trainees’ interactions both inside and out of the hospital setting. However, there were concerns about the quality of teaching and team-members’ interactions, especially when the cameras were not used. More studies are needed to explore if the virtual part of the clinical round is best suited for the patients’ management and HCWs’ experience.

## Data Availability

All the data for this study will be made available upon reasonable request.

## Abbreviations

CDC: Centers for Disease Control and Prevention
COVID-19: coronavirus disease 2019
ePPE: electronic personal protective equipment
KSUMC: King Saud University Medical City
PICU: Pediatric Intensive care unit
WHO: World Health Organization

## Acknowledgment

We are grateful to all the PICU team and all HCWs on the front lines of COVID-19. We are also thankful to Consulla Data analysis and statistical consulting for their support.

## Appendix 1: Details of the PICU hybrid-style round

### RE: Hybrid-style Zoom for in-hospital PICU video-telemedicine

Our PICU will pilot Hybrid-style daily rounds, to optimize Social Distancing recommendations for COVID prevention, while ensuring proper utilization of PICU resources to what the patient’s needs, while maintaining academic bedside teachings for the rotating residents and fellows.

- <u>All the designated PICU team members must be inside the hospital at the time of this hybrid-style PICU rounds,</u> but in separates physical locations during the thorough discussions of each patient’s condition, to promote physical distancing.
- The senior PICU physician should initiate the Zoom meeting number (with password) and share it with the other PICU Team members (physicians, nurses, RT, pharmacist, dietician).
- All PICU Team members on that day are expected to join this endorsement within 5 minutes of the starting time. Video should be on, to optimize the 2-ways communications, like the current hand-over process.
- Please ensure that you know all the colleagues who are attending your videoconference before discussing any managements for your patients. <u>No recordings on Zoom, to maximize patient’s privacy.</u>

### The steps for the Hybrid Round are

1. Before the Zoom meeting starts, the PICU physicians will each examine, assess and stabilize their patient like the conventional style, before the hybrid rounds starts.
2. The rotating resident will present her/his PICU patient, with backup and clarifications from the designated PICU seniors. Input from the multidisciplinary team is encouraged for each patient.
3. After this thorough Zoom discussion of all PICU patients, the PICU Consultant will make physical rounds in the PICU, to complement any further details and ensure the management plan and family counselling are done.
4. The rotating residents are encouraged to actively engage in the management plan and decision making, as appropriate to their learning objectives
  - If there is any technical difficulties and the meeting could not be resolved within 5 minutes, to switch back to the conventional PICU daily round process, making sure that all comply with the hospital’s Infection Control measures.
  - to ensure accurate documentation, as well as full patient’s privacy, the process and recommendations must be documented in the patient’s EHR as per the hospital policy. The PICU Consultant will counter-sign these documentation in the EHR.
  - This will be piloted in the first month, to ensure the verbatim component and clear communication that were done through the Zoom.
  - By the end of the one-month pilot, a report on feasibility and effectiveness with feedback from the PICU team, will be sent from the PICU Head to the Pediatric Chairman.

**Stay safe!**

6-9-2020

## Appendix 2: The focused group themes and questions

Hybrid PICU round:

Focused group themes and questions:

Overall experience:

- How do you think was the hybrid PICU round experience?
- What were the aims of this hybrid style PICU round?
- How much did we achieve of these aims?
- What do you think about the starting time of the Zoom hybrid round?
- What do you think about the duration of the Zoom and the physical rounds?
- What do you think about the structure of the Hybrid round?
- Should the bedside physical round be before or after the Zoom round?
- Why? What do you think about the attendees during the hybrid PICU round? How should attend? How should not?

Facilitating factors:

- From your actual experience in the hybrid PICU round, what things made it more fruitful?
- Share with us some examples…

Opposing factors:

- From your actual experience in the hybrid PICU round, what things made it more difficult?
- Share with us some examples…

Multidisciplinary Team interactions:

- How were the multidisciplinary Team discussions during the hybrid round as compared to before it?

Patients’ care:

- What were the effects of the Hybrid PICU round on the patient’s care during the pandemic?
- How can it be improved?

Teachings in the round:

- What were the effects of the Hybrid PICU round on the educational process?
- How can it be improved?

Conclusion:

- What other clinical areas can utilize a similar hybrid round style?
- How can we improve the current hybrid round?
- Shall we continue this hybrid style PICU round after the pandemic? Why?

## Ref

1. W. H. Organization: WHO Coronavirus (COVID-19) Dashboard. Available at: https://covid19.who.int/. Accessed April 18 2021,

2. L. The: COVID-19: protecting healthcare workers,. Lancet 2020; 395:922

3. L. H. Nguyen, D. A. Drew, M. S. Graham, A. D. Joshi, C. G. Guo, W. Ma, R. S. Mehta, E. T. Warner, D. R. Sikavi, C. H. Lo, S. Kwon, M. Song, L. A. Mucci, M. J. Stampfer, W. C. Willett, A. H. Eliassen, J. E. Hart, J. E. Chavarro, J. W. Rich-Edwards, R. Davies, J. Capdevila, K. A. Lee, M. N. Lochlainn, T. Varsavsky, C. H. Sudre, M. J. Cardoso, J. Wolf, T. D. Spector, S. Ourselin, C. J. Steves and A. T. Chan: Risk of COVID-19 among front-line healthcare workers and the general community: a prospective cohort study. Lancet Public Health 2020; 5:e475–e483

4. L. Ling, W. T. Wong, W. T. P. Wan, G. Choi and G. M. Joynt: Infection control in non-clinical areas during the COVID-19 pandemic. Anaesthesia 2020; 75:962–963

5. C. Stickrath, M. Noble, A. Prochazka, M. Anderson, M. Griffiths, J. Manheim, S. Sillau and E. Aagaard: Attending Rounds in the Current Era: What Is and Is Not Happening. JAMA Internal Medicine 2013; 173:1084–1089

6. R. P. Dutton, C. Cooper, A. Jones, S. Leone, M. E. Kramer and T. M. Scalea: Daily multidisciplinary rounds shorten length of stay for trauma patients. Journal of Trauma and Acute Care Surgery 2003; 55:913–919

7. M.-H. Temsah, N. Abouammoh, A. Ashry, A. Al-Eyadhy, A. Alhaboob, F. Alsohime, M. Almazyad, M. Alabdulhafid, R. Temsah and F. Aljamaan: Virtual handover of patients in the pediatric intensive care unit during COVID-19 crisis. medRxiv 2021

8. CDC: COVID-19 in a Correctional Facility Employee Following Multiple Brief Exposures to Persons with COVID-19 — Vermont, July–August 2020. Available at: https://www.cdc.gov/mmwr/volumes/69/wr/mm6943e1.htm. Accessed April 18 2021,

9. R. Abdelmoniem, R. Fouad, S. Shawky, K. Amer, T. Elnagdy, W. A. Hassan, A. M. Ali, M. Ezzelarab, Y. Gaber, H. A. Badary, S. Musa, H. Talaat, A. M. Kassem and O. Tantawi: SARS-CoV-2 infection among asymptomatic healthcare workers of the emergency department in a tertiary care facility. Journal of Clinical Virology 2021; 134:104710

10. D. R. Garfin: Technology as a coping tool during the coronavirus disease 2019 (COVID-19) pandemic: Implications and recommendations. Stress and Health 2020; 36:555–559

11. S. Chandra, C. Hertz, H. Khurana and M. E. Doerfler: Collaboration Between Tele-ICU Programs Has the Potential to Rapidly Increase the Availability of Critical Care Physicians-Our Experience Was During Coronavirus Disease 2019 Nomenclature. Critical care explorations 2021; 3:e0363–e0363

12. R. W. Turer, I. Jones, S. T. Rosenbloom, C. Slovis and M. J. Ward: Electronic personal protective equipment: A strategy to protect emergency department providers in the age of COVID-19. Journal of the American Medical Informatics Association 2020; 27:967–971

13. A. Hagana, R. Behranwala, N. Aojula and N. Houbby: Digitalising medical education: virtual ward rounds during COVID-19 and beyond. BMJ Simulation and Technology Enhanced Learning 2020:bmjstel-2020-000742

14. D. Vogel and S. Harendza: Basic practical skills teaching and learning in undergraduate medical education - a review on methodological evidence. GMS J Med Educ 2016; 33:Doc64

15. H. Hofmann, C. Harding, J. Youm and W. Wiechmann: Virtual bedside teaching rounds with patients with COVID-19. Med Educ 2020; 54:959–960

16. R. J. Wilcha: Effectiveness of Virtual Medical Teaching During the COVID-19 Crisis: Systematic Review. JMIR Med Educ 2020; 6:e20963

17. S. M. Badawy and A. Radovic: Digital Approaches to Remote Pediatric Health Care Delivery During the COVID-19 Pandemic: Existing Evidence and a Call for Further Research. JMIR Pediatr Parent 2020; 3:e20049

18. V. J. Hall, S. Foulkes, A. Saei, N. Andrews, B. Oguti, A. Charlett, E. Wellington, J. Stowe, N. Gillson, A. Atti, J. Islam, I. Karagiannis, K. Munro, J. Khawam, M. A. Chand, C. S. Brown, M. Ramsay, J. Lopez-Bernal and S. Hopkins: COVID-19 vaccine coverage in healthcare workers in England and effectiveness of BNT162b2 mRNA vaccine against infection (SIREN): a prospective, multicentre, cohort study. Lancet 2021; 397:1725–1735

19. M. G. Thompson, J. L. Burgess, A. L. Naleway, H. L. Tyner, S. K. Yoon, J. Meece, L. E. W. Olsho, A. J. Caban-Martinez, A. Fowlkes, K. Lutrick, J. L. Kuntz, K. Dunnigan, M. J. Odean, K. T. Hegmann, E. Stefanski, L. J. Edwards, N. Schaefer-Solle, L. Grant, K. Ellingson, H. C. Groom, T. Zunie, M. S. Thiese, L. Ivacic, M. G. Wesley, J. M. Lamberte, X. Sun, M. E. Smith, A. L. Phillips, K. D. Groover, Y. M. Yoo, J. Gerald, R. T. Brown, M. K. Herring, G. Joseph, S. Beitel, T. C. Morrill, J. Mak, P. Rivers, K. M. Harris, D. R. Hunt, M. L. Arvay, P. Kutty, A. M. Fry and M. Gaglani: Interim Estimates of Vaccine Effectiveness of BNT162b2 and mRNA-1273 COVID-19 Vaccines in Preventing SARS-CoV-2 Infection Among Health Care Personnel, First Responders, and Other Essential and Frontline Workers - Eight U.S. Locations, December 2020-March 2021. MMWR Morb Mortal Wkly Rep 2021; 70:495–500

20. E. Hacisuleyman, C. Hale, Y. Saito, N. E. Blachere, M. Bergh, E. G. Conlon, D. J. Schaefer-Babajew, J. DaSilva, F. Muecksch, C. Gaebler, R. Lifton, M. C. Nussenzweig, T. Hatziioannou, P. D. Bieniasz and R. B. Darnell: Vaccine Breakthrough Infections with SARS-CoV-2 Variants. N Engl J Med 2021

21. A. M. Cavanaugh, S. Fortier, P. Lewis, V. Arora, M. Johnson, K. George, J. Tobias, S. Lunn, T. Miller, D. Thoroughman and K. B. Spicer: COVID-19 Outbreak Associated with a SARS-CoV-2 R.1 Lineage Variant in a Skilled Nursing Facility After Vaccination Program - Kentucky, March 2021. MMWR Morb Mortal Wkly Rep 2021; 70:639–643

22. R. A. Teran, K. A. Walblay, E. L. Shane, S. Xydis, S. Gretsch, A. Gagner, U. Samala, H. Choi, C. Zelinski and S. R. Black: Postvaccination SARS-CoV-2 Infections Among Skilled Nursing Facility Residents and Staff Members - Chicago, Illinois, December 2020-March 2021. MMWR Morb Mortal Wkly Rep 2021; 70:632–638

23. CDC: Interim Public Health Recommendations for Fully Vaccinated People. Available at: https://www.cdc.gov/coronavirus/2019-ncov/vaccines/fully-vaccinated-guidance.html#print. Accessed May 30 2021,

24. S. Thusini: Critical care nursing during the COVID-19 pandemic: a story of resilience. Br J Nurs 2020; 29:1232–1236

25. M. Vitacca, M. Carone, E. M. Clini, M. Paneroni, M. Lazzeri, A. Lanza, E. Privitera, F. Pasqua, F. Gigliotti, G. Castellana, P. Banfi, E. Guffanti, P. Santus and N. Ambrosino: Joint Statement on the Role of Respiratory Rehabilitation in the COVID-19 Crisis: The Italian Position Paper. Respiration 2020; 99:493–499

26. K. J. Roberts, B. Johnson, H. M. Morgan, J. M. Vrontisis, K. M. Young, E. Czerpak, B. D. Fuchs and M. Pierce: Evaluation of Respiratory Therapist Extender Comfort With Mechanical Ventilation During COVID-19 Pandemic. Respir Care 2021; 66:199–204

27. C. H. Li, A. G. Rajamohan, P. T. Acharya, C. J. Liu, V. Patel, J. L. Go, P. E. Kim and J. Acharya: Virtual Read-Out: Radiology Education for the 21st Century During the COVID-19 Pandemic. Acad Radiol 2020; 27:872–881

28. E. S. Yuan: End-to-End Encryption Update. Zoom Blog. URL: https://blog.zoom.us/end-to-end-encryption-update/ [Accessed 2020-07-26] 2020

29. M. Akhtar, P. G. Van Heukelom, A. Ahmed, R. D. Tranter, E. White, N. Shekem, D. Walz, C. Fairfield, J. P. Vakkalanka and N. M. Mohr: Telemedicine Physical Examination Utilizing a Consumer Device Demonstrates Poor Concordance with In-Person Physical Examination in Emergency Department Patients with Sore Throat: A Prospective Blinded Study. Telemed J E Health 2018; 24:790–796

30. A. M. Ansary, J. N. Martinez and J. D. Scott: The virtual physical exam in the 21st century. J Telemed Telecare 2019:1357633x19878330

31. C. P. Benziger, M. D. Huffman, R. N. Sweis and N. J. Stone: The Telehealth Ten: A Guide for a Patient-Assisted Virtual Physical Examination. Am J Med 2021; 134:48–51

32. O. Jumreornvong, E. Yang, J. Race and J. Appel: Telemedicine and Medical Education in the Age of COVID-19. Acad Med 2020; 95:1838–1843

33. K. Lawrence, K. Hanley, J. Adams, D. J. Sartori, R. Greene and S. Zabar: Building Telemedicine Capacity for Trainees During the Novel Coronavirus Outbreak: a Case Study and Lessons Learned. J Gen Intern Med 2020; 35:2675–2679

34. D. A. Fleming, S. L. Riley, S. Boren, K. G. Hoffman, K. E. Edison and C. S. Brooks: Incorporating telehealth into primary care resident outpatient training. Telemed J E Health 2009; 15:277–282

35. M. S. Lee and V. Nambudiri: Integrating Telemedicine Into Training: Adding Value to Graduate Medical Education Through Electronic Consultations. J Grad Med Educ 2019; 11:251–254

36. M. J. Grzyb, H. Coo, L. Rühland and K. Dow: Views of parents and healthcare providers regarding parental presence at bedside rounds in a neonatal intensive care unit. J Perinatol 2014; 34:143–148

37. T. C. Ingram, P. Kamat, C. M. Coopersmith and A. Vats: Intensivist perceptions of family-centered rounds and its impact on physician comfort, staff involvement, teaching, and efficiency. J Crit Care 2014; 29:915–918

38. P. R. Gupta, R. S. Perkins, R. L. Hascall, C. F. Shelak, S. Demirel and M. T. Buchholz: The Effect of Family Presence on Rounding Duration in the PICU. Hosp Pediatr 2017; 7:103–107

39. J. Gaulton, K. Ziegler and E. Chang: Virtual Practices Transform the Care Delivery Model in an Intensive Care Unit During the Coronavirus Pandemic. Nejm Catalyst Innovations in Care Delivery 2020:10.1056/CAT.1020.0169

40. N. M. Howard, D. A. Cook, R. Hatala and M. V. Pusic: Learning Curves in Health Professions Education Simulation Research: A Systematic Review. Simul Healthc 2021; 16:128–135

